# Adaptation of the Person-Centered Maternity Care scale for people of color in the United States

**DOI:** 10.1101/2021.05.06.21256758

**Authors:** Patience A. Afulani, Molly R. Altman, Esperanza Castillo, Nayeli Bernal, Linda Jones, Tanefer Camara, Zoe Carrasco, Shanell Williams, May Sudhinaraset, Miriam Kuppermann

**Affiliations:** Department of Epidemiology and Biostatistics, University of California; Department of Obstetrics, Gynecology, & Reproductive Sciences, University of California, San Francisco; Department of Child, Family, and Population Health Nursing, University of Washington; California Preterm Birth Initiative, University of California, San Francisco; Department of Community Health Science, University of California, Los Angeles

**Keywords:** Person-centered care, maternity care, intrapartum care, birth, people of color, Black women, community-based participatory research, respectful maternity care, experience of care, quality of care, measurement

## Abstract

**Introduction:** Mistreatment by healthcare providers disproportionately affects people of color in the United States (US). The goal of this study is to adapt the global Person-Centered Maternity Care (PCMC) scale to the experiences of people of color in the US using a community-engaged approach.

**Methods:** We conducted expert reviews to improve content validity and cognitive interviews with potential respondents were conducted to assess relevance, comprehension, and comprehensiveness. Surveys of 297 postpartum people, 82% of whom identified as Black, were used for psychometric analysis in which we assessed construct and criterion validity and reliability. The University of California, San Francisco, California Preterm Birth Initiative’s Community Advisory Board (CAB), which consists of community members, community-based health workers, and social service providers in Northern California, provided input during all stages of the project.

**Results:** Through an iterative process of factor analysis, discussions with the CAB, and a prioritization survey, we eliminated items that performed poorly in psychometric analysis, yielding a 35-item PCMC-US scale with sub-scales for “dignity and respect,” “communication and autonomy,” and “responsive and supportive care.” The Cronbach’s alpha for the full scale is 0.95 and for the sub-scales is 0.87. Standardized summative scores range from 0 to 100, with higher scores indicating higher PCMC. Correlations with related measures indicated high criterion validity.

**Conclusions:** The 35-item PCMC-US scale and its sub-scales have high validity and reliability in a sample of predominantly Black women. This scale provides a tool to support efforts to reduce the disparities in birth outcomes among people of color.

## INTRODUCTION

Disrespectful care and mistreatment by healthcare providers have been shown to disproportionately affect people of color in the United States (US), particularly Black pregnant and birthing people, and have been attributed to racism and discrimination (Alhusen et al., 2016; Altman et al., 2019; McLemore et al., 2018; Vedam, Stoll, Taiwo, et al., 2019). Hospitalizations for labor and delivery are often considered the apex for these experiences, not only because birth is a major life transition, but also due to exposure to unfamiliar providers and a culture of care that may or may not be aligned with patient expectations (Lyndon et al., 2018).

Person-centered care, or care that emphasizes the wants and needs of the patient over the provider, focuses on respect, trust, dignity, support, autonomy, and communication among other domains, and represents a major determinant of high-quality care provision (Institute of Medicine (US) Committee on Quality of Health Care in America, 2001; WHO, 2016). Various aspects of person-centered care during childbirth are affected by implicit bias and are often dependent on providers’ assumptions regarding the ability of birthing people to make decisions about their care (Afulani, Ogolla, et al., 2021; Altman et al., 2019; Vedam, Stoll, McRae, et al., 2019).

While numerous qualitative studies have examined aspects of person-centered maternity care, very few have attempted to operationalize it quantitatively to enable measurement of the effectiveness of interventions aimed at improving quality of care, and, to our knowledge, none have looked at the unique experiences of Black pregnant and birthing people in the US (Nilvér et al., 2017). We sought to adapt and validate a scale to measure person-centered care during labor and childbirth to reflect the experiences of people of color in the US, with an emphasis on the experiences of Black women and birthing people.

## MATERIALS AND METHODS

The Person-Centered Maternity Care (PCMC) scale, which was initially developed for use in low resource countries and has been validated in several contexts (Afulani et al., 2017, 2018; Afulani, Phillips, et al., 2019), served as the basis for this US-focused version. The original PCMC scale was first validated in Kenya and subsequently in India and Ghana (Afulani et al., 2018; Afulani, Phillips, et al., 2019; Afulani, Aborigo, et al., 2019). To adapt this scale for use in the US, we followed standard scale development procedures in addition to a community-engaged approach to ensure that it is valid, reliable, and fits the needs of Black birthing people and families (DeVellis, 2016; Hinkin et al., 1997). Specifically, a literature review was conducted to define the construct and identify domains and to develop the initial list of items (Afulani et al., 2017). We then adapted the scale using expert reviews, cognitive interviews, structured surveys, and psychometric analysis—engaging community members throughout the process—and named the adapted version the PCMC-US scale. We used a parallel process to develop a prenatal care-oriented measure, which we refer to as the Person-Centered Prenatal Care (PCPC) scale (Afulani, Altman, et al., 2021).

### Expert reviews

The goal of the expert review was to optimize content validity in terms of relevance and comprehensiveness (DeVellis, 2016). Our research team, which included clinicians and researchers, first reviewed the 30 items in the original PCMC scale and removed items not applicable to the US setting (e.g., availability of water and electricity). We then held an expert review session with 10 members of the University of California, San Francisco (UCSF), California Preterm Birth Initiative’s (CA-PTBi) Community Advisory Board (CAB). The CAB includes people of color who have had preterm births, community-based health workers, and social service providers in Northern California. The CAB members represent community experts who understand what PCMC means to people of color because of their direct service and lived experiences in the communities of focus. A second expert review session consisted of 20 people, including two community health workers, four CAB members, and 14 faculty members (researchers and clinicians). During these sessions, the remaining questions were reviewed, and several new questions were recommended that related to existing PCMC domains (Examples: “Did you feel heard and listened to?” “Did providers encourage you to ask questions?” “Did providers knock on your room’s door and wait for response before entering?”). Questions on infant feeding choices and emotional well-being were also added. CAB members were paid $40/hour for their participation in these sessions.

### Cognitive interviews

We next conducted cognitive interviews to assess and improve comprehensibility, relevance, and comprehensiveness of the questions (Collins, 2003; Nápoles-Springer et al., 2006). We developed an interview guide that included questions on item wording, possible sources of confusion, and appropriateness of items, as well as whether questions were missing. We then conducted the cognitive interviews with 15 participants who were ≥28 weeks pregnant or had given birth within the past year. Participants were recruited from partner organizations and from a UCSF database of patients who had previously participated in studies and were interested in being contacted for future studies. Participants received a $50 gift card for participation. Feedback from the cognitive interviews were used to revise the items. We had 50 candidate items for the PCMC-US scale (Table 1) at the end of the cognitive interviews. These questions were then integrated into the study questionnaire that included items on sociodemographic characteristics and other issues. The questionnaire was piloted with eight people meeting eligibility criteria for the planned survey to assure acceptability and ease of completion.

**Table 1:**
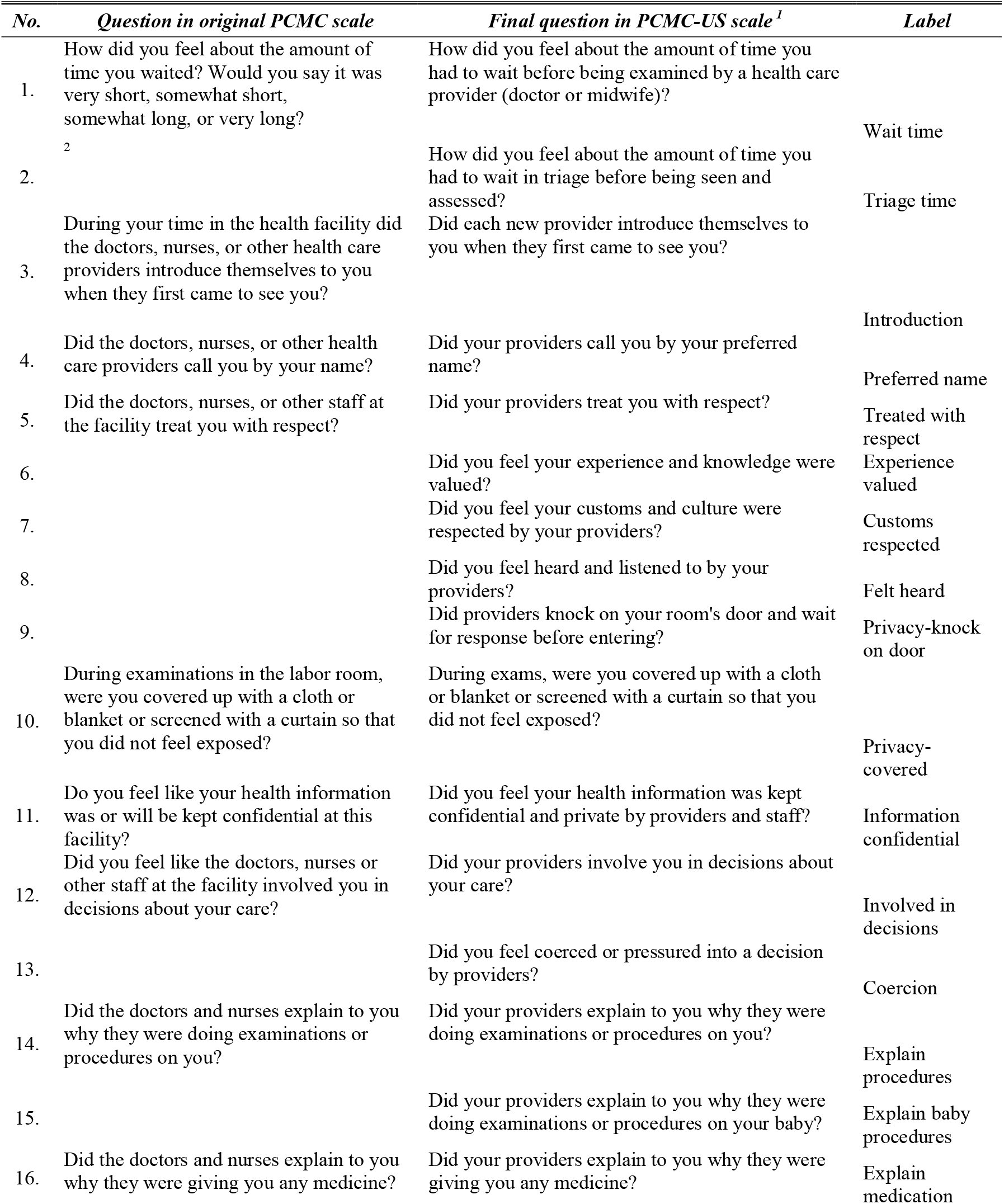
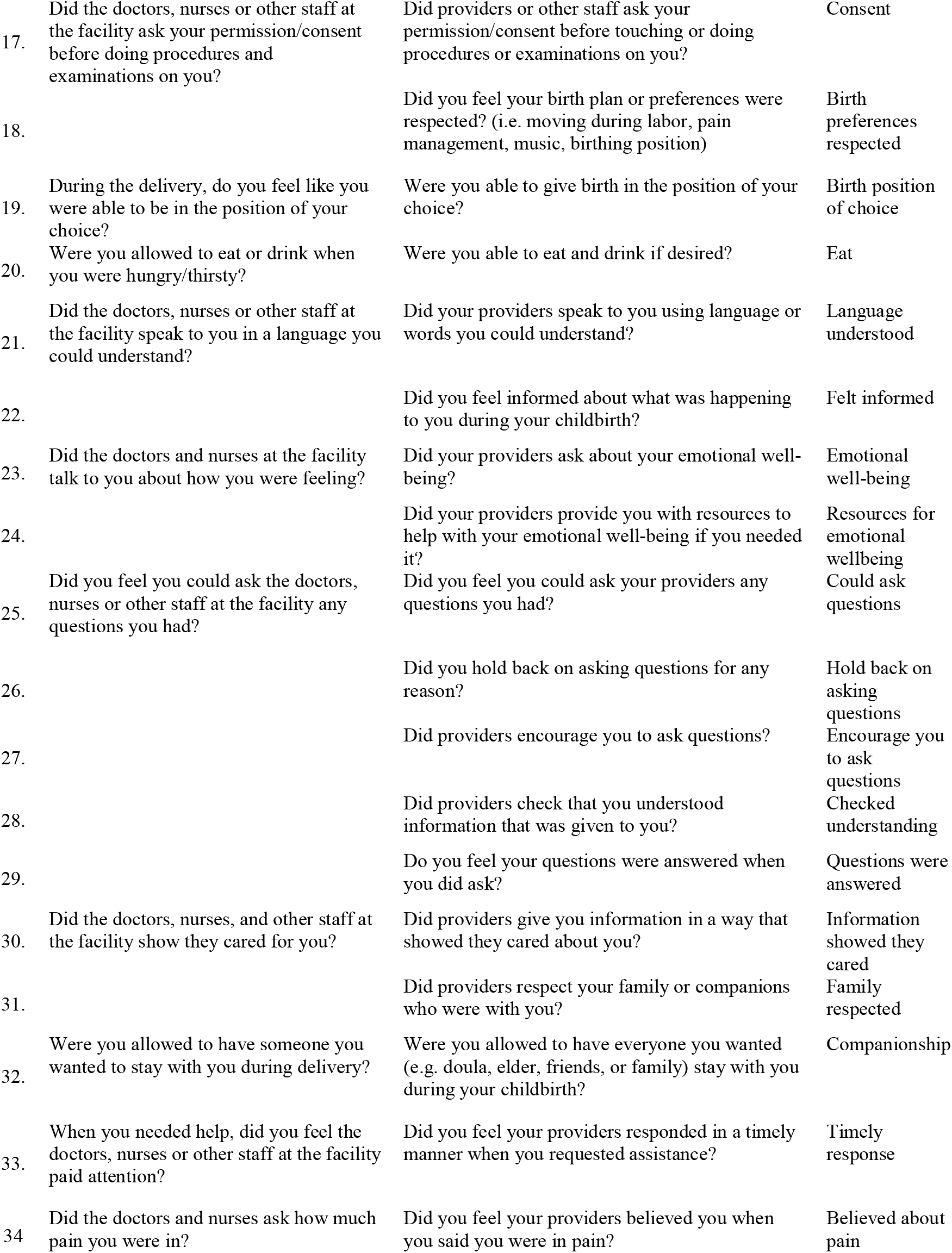
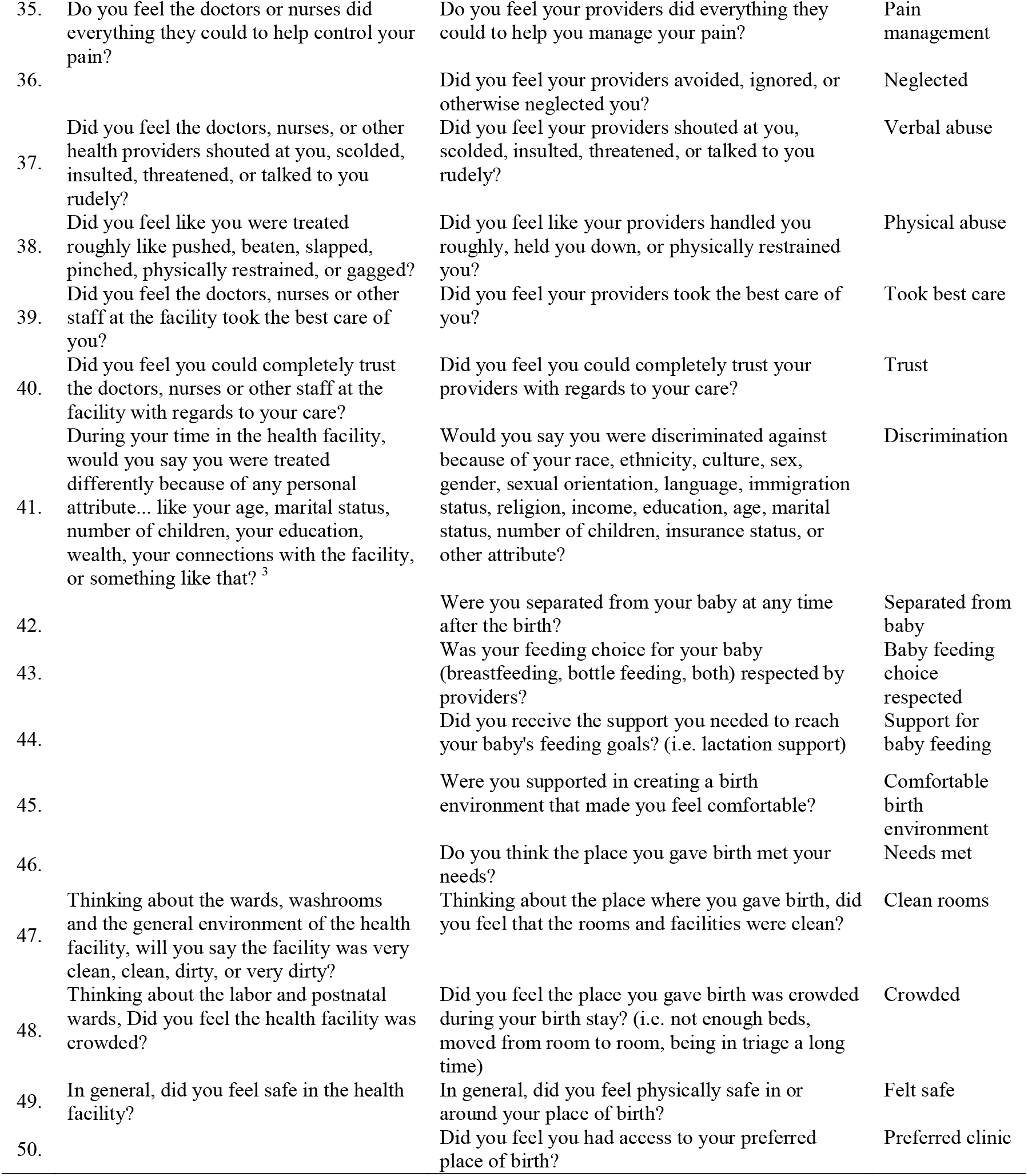
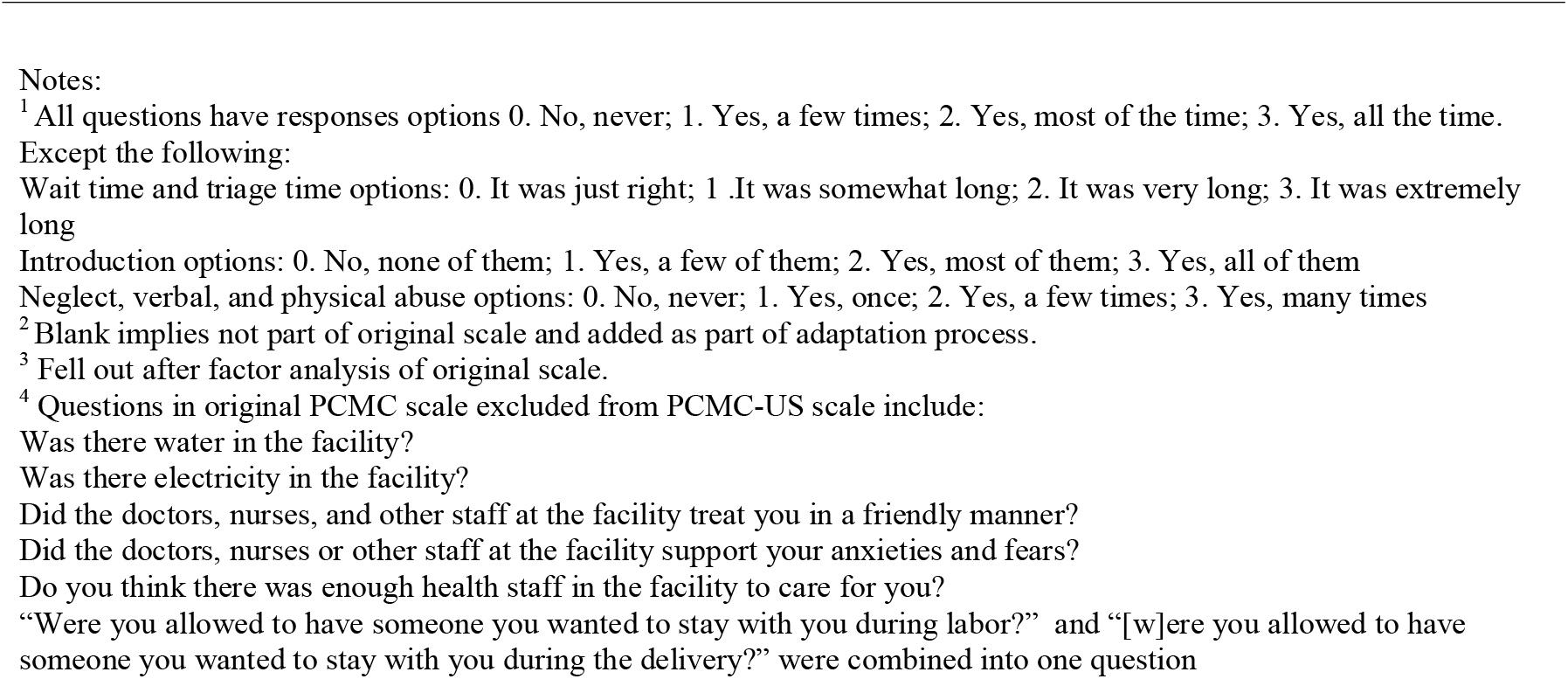
Person-Centered Maternity Care (PCMC) questions.

### Survey

Participants for the survey were recruited through community-based organizations and targeted social media advertisements. Individuals aged 15 years or older who had given birth in the past year were eligible to participate, and recruitment materials indicated that the study was focused on the experiences of people of color. We conducted the survey between January and September 2020 using the REDCap (Research Electronic Database Capture) online survey platform for data collection (Harris et al., 2009). Individuals who were interested in joining the study emailed the study team, answered eligibility questions, and once confirmed as eligible, were emailed a personalized link to complete the survey. Study information was provided on the landing page and respondents clicked a button to indicate informed consent before the survey opened up. Participants received a $40 electronic gift card upon completing the survey. Ethical approval was obtained from UCSF’s Institutional Review Board.

### Psychometric analyses

The goal of the psychometric analyses was to assess and improve construct and criterion validity and internal reliability of the scale (DeVellis, 2016; Hinkin et al., 1997). We examined the distributions of all the items and recoded some response options to obtain a uniform scale. To ensure that all response options ranged from 0 to 3, we recoded items that had a “not applicable” category (4) to the upper middle category (2: most of the time). We also reverse-coded negative items so that all responses reflected a scale ranging from 0 as the lowest level to 3 as the highest level. We then constructed a correlation matrix to examine the correlations among the items.

Construct validity was assessed using exploratory factor analysis (DeVellis, 2016; Hinkin et al., 1997). The Kaiser-Meyer-Olkin (KMO) measure was used to test suitability of the items for factor analysis. The decision on how many factors to retain was made using Kaiser’s rule of retaining only factors with eigenvalues ≥1, and examining the “elbow” of the scree plot (plot of eigenvalues) (Afifi et al., 2004; DeVellis, 2016; Hinkin et al., 1997). Oblique rotation, which allows for correlation between the rotated factors, was used because the person-centered care domains are theoretically related (Afifi et al., 2004; Hinkin et al., 1997). We used initial criteria of factor loadings ≥0.3 and uniqueness of ≤0.8 in iterative factor analysis, combined with feedback from the CAB to determine which items to retain or exclude (Afifi et al., 2004). Sub-scales were generated using the loading of the items on the extracted factors and conceptual groupings, and the items were summed to generate the PCMC-US scale and sub-scale scores.

Internal consistency reliability was assessed using Cronbach’s alpha (DeVellis, 2016; Hinkin et al., 1997). To assess criterion validity, we examined whether scales and sub-scales were associated with theoretically related measures (DeVellis, 2016). For example, we hypothesized that the PCMC-US scale would be associated with satisfaction and perceptions of the quality of intrapartum care received, and whether the participant would use the same service in the future (Afulani et al., 2017; Sheferaw et al., 2016). We also examined the association between the PCMC-US scale and two other measures which assess women’s combined experiences during pregnancy and childbirth: the Mothers Autonomy in Decision Making scale (MADM) and the Mothers on Respect Index (MORi) index (Vedam, Stoll, Martin, et al., 2017; Vedam, Stoll, Rubashkin, et al., 2017). We tested our hypotheses through correlations and bivariate linear and logistic regression analysis. STATA version 15 was used for all analyses.

## RESULTS

In all, 312 participants completed the study. Fifteen participants who did not respond to all the PCMC-US items were excluded, resulting in an analytic sample of 297. Most respondents identified as Black (82%). The average age was 29 years old (SD=3.6), and most participants were married (89%), primiparous (76%), and college graduates (52%; Table 2).

**Table 2:** Characteristics of respondents (N=297)

|  | No. | % |
| --- | --- | --- |
| Race/ethnicity |  |  |
| Black/African American | 242 | 81.5 |
| White/Caucasian | 33 | 11.1 |
| Asian | 5 | 1.7 |
| Hawaiian/Pacific Islander | 3 | 1.0 |
| Latina/Hispanic | 18 | 6.1 |
| American Indian/Alaskan Native | 3 | 1.0 |
| Multiracial | 2 | 0.7 |
| Other | 5 | 1.7 |
| Prefer not to answer | 3 | 1.0 |
| Partner's race/ethnicity |  |  |
| Black/African American | 240 | 80.8 |
| White/Caucasian | 35 | 11.8 |
| Asian | 4 | 1.3 |
| Hawaiian/Pacific Islander | 1 | 0.3 |
| Latinx/Hispanic | 18 | 6.1 |
| American Indian/Alaskan Native | 2 | 0.7 |
| Multiracial | 1 | 0.3 |
| Other | 3 | 1.0 |
| Prefer not to answer | 2 | 0.7 |
| No partner | 2 | 0.7 |
| Age |  |  |
| 17-28 | 102 | 34.3 |
| 29-32 | 125 | 42.1 |
| 33-45 | 70 | 23.6 |
| Married | 263 | 88.6 |
| Primiparous | 225 | 75.8 |
| Time since delivery |  |  |
| <3 months | 103 | 34.7 |
| 3-4 months | 106 | 35.7 |
| 5-12 months | 88 | 29.6 |
| Educational attainment |  |  |
| High school or less | 38 | 12.8 |
| Some college | 106 | 35.7 |
| College graduate | 153 | 51.5 |
| Average yearly income |  |  |
| <\$50,000 | 62 | 20.9 |
| \$50,0001-\$100,000 | 185 | 62.3 |
| >\$100,000 | 50 | 16.8 |
| Employment status |  |  |
| Not employed | 104 | 35.0 |
| Employed full-time | 152 | 51.2 |
| Employed part-time | 37 | 12.5 |
| Prefer not to answer | 4 | 1.3 |
| Insurance type |  |  |
| No insurance | 7 | 2.4 |
| Private/employer-provided insurance | 243 | 81.8 |
| Medicaid/MediCal | 16 | 5.4 |
| Medicare | 22 | 7.4 |
| Tricare/government | 2 | 0.7 |
| Prefer not to answer | 7 | 2.4 |

The distributions of the 50 individual PCMC-US items are in Appendix A. With few exceptions, the responses ranged between 0 and 3. The proportion of N/A responses on items that generated N/A responses ranged from 1 to 13% except for the question regarding whether providers explained medications, for which 54% responded that they did not receive any medicine. There was good correlation between most items, which generally ranged from 0.2 to 0.7. Items that had correlations of <2 or negative correlations with other items were flagged. These included the question on preferred name, preferred language, position of choice, being separated from the baby, support for feeding goals, cleanliness, coercion, holding back from asking questions, and crowding.

The KMO measures of sampling adequacy for all items ranged from 0.69 to 0.97, with an overall KMO of 0.94, indicating suitability for factor analysis. The initial factor analysis with all 50 items yielded 6 factors with eigenvalues of ≥1, accounting for 84% of the cumulative variance. However, there was one dominant factor (Figure 1) with an eigenvalue of 16.8, which accounted for 60% of cumulative variance. The eigenvalues for the other factors were <2 (range: 1.0 to 1.6). All the items had a factor loading of ≥0.3 on at least one of the 6 factors except the questions on introduction, language, informed about what was happening, and support for feeding goals, which had loadings of between 0.2 and 0.3. Uniqueness for all items was <0.8 except for introduction, separation, language, holding back from asking questions, and crowding, which had uniqueness between 0.8 and 0.9. When the analysis was restricted to the single factor structure, all items had loadings of >0.3 except the items on name, separation, explained medications, coercion, holding back from asking questions, and crowding, which had loadings of <0.2 and uniqueness >0.9. We conducted iterative factor analysis dropping different items and examining the results. Given the need for a parsimonious scale, we also identified items that were conceptually similar for possible elimination.

**Figure 1:**
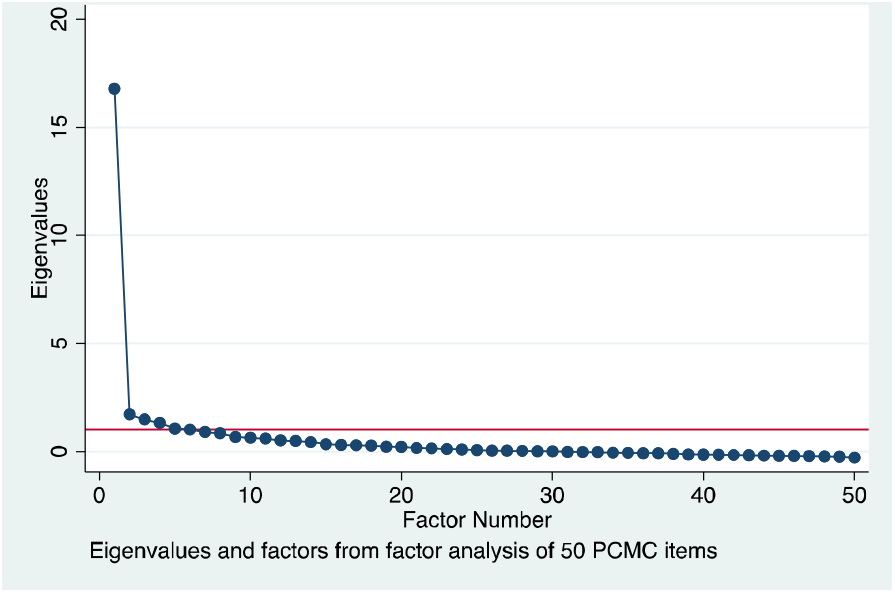
Scree plot from factor analysis of 50 PCMC items

We presented the initial findings to the CAB and suggested dropping the items that had low loadings, high uniqueness, or conceptual difference/similarity. Although some CAB members acknowledged 50 items were too many for a scale, many strongly felt that all questions were important and should be included, so we could not achieve consensus on items to drop. Thus, to enable us to get to a smaller subset of questions, we used the results of the factor analysis, as well as conceptual domains based on the World Health Organization experience of care domains (Tunçalp et al., 2015), to group the 50 items into three domains for “Dignity and Respect” (DR: 11 items), “Communication and Autonomy” (CA: 19 items), and “Responsive and Supportive Care” (RS: 20 items) We then created a survey, grouping conceptually similar items together, and asked CAB members as well as research team members to select their top 8 to 10 questions from each domain.

The results of this survey (completed by 14 people and shown in Appendix B) were combined with the factor analysis to select items for the scale. We first decided to drop all items selected by less than a third of respondents (<5), as this suggested low priority, resulting in the elimination of 13 items (4 from CA, 1 from DR, and 8 from RS). We then re-ran the factor analysis on the remaining 37 items. All but three items had loadings >0.3 on the single factor for each domain and on the full scale (coercion, called by preferred name, and being separated from baby). Since the items on preferred name and being separated from baby were not highly ranked in the survey (selected by only 5 people), we decided to drop these two items next, but retained the coercion item despite its low loading because it was recommended by most respondents (9) and was one of the items CAB members felt strongly about retaining. Of note, despite reporting a generally good experience, about 30% of participants in this sample reported being pressured into a decision by their providers, pointing to the importance of this item. This yielded a 35-item scale with 3 factors (Figure 2). All items had loadings >0.3 on the full scale and on the single factor for each domain, except the item on coercion (Table 3). Dropping the coercion item yielded a 34-item version in which all had loadings >0.3 on the full scale and on the single factor for each domain. Inclusion of the coercion item, however, did not significantly change the loadings of the other items or the Cronbach’s alpha (which was 0.95 in both cases). So the 35-item version was maintained with 14, 10, and 11 items respectively for the CA, DR, and RS sub-scales. The Cronbach’s alpha for all the sub-scales is 0.87 (Table 4).

**Figure 2:**
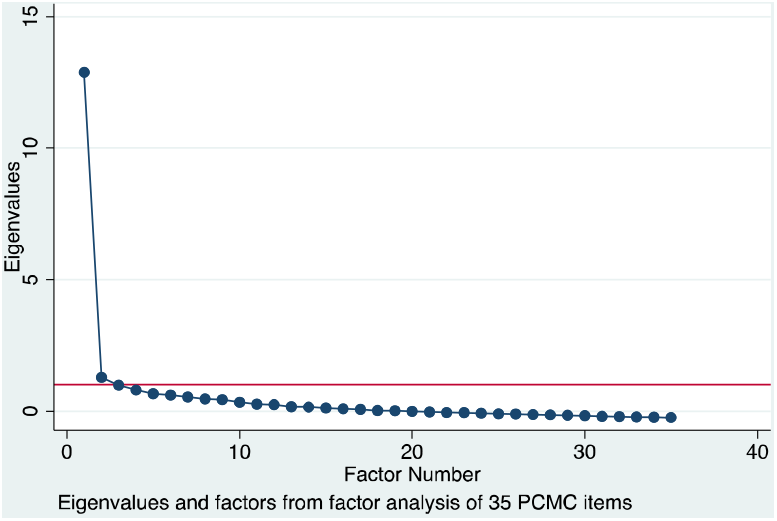
Scree plot from factor analysis of 35 PCMC items.

**Table 3:** Results of exploratory factor analysis for 35-item Person-Centered Maternity Care – US scale (N=297)

|  | Single factor |  | Three factor structure |  |  |  | Loading on individual sub-scales |  |  |  |
| --- | --- | --- | --- | --- | --- | --- | --- | --- | --- | --- |
|  | F1 | Uniqueness | F1 | F2 | F3 | Uniqueness | CA | DR | RS | Uniqueness |
| 1. Introduction | 0.36 | 0.87 |  | 0.23 |  | 0.86 | 0.36 |  |  | 0.87 |
| 2. Felt heard | 0.72 | 0.49 | 0.33 | 0.47 |  | 0.47 | 0.75 |  |  | 0.44 |
| 3. Involved in decisions | 0.77 | 0.41 | 0.45 | 0.42 |  | 0.39 | 0.79 |  |  | 0.37 |
| 4. Explain procedures | 0.68 | 0.54 |  | 0.59 |  | 0.47 | 0.72 |  |  | 0.49 |
| 5. Consent | 0.59 | 0.66 |  | 0.69 |  | 0.55 | 0.62 |  |  | 0.62 |
| 6. Language understood | 0.40 | 0.84 | 0.28 |  |  | 0.82 | 0.48 |  |  | 0.77 |
| 7. Felt informed | 0.55 | 0.70 |  | 0.44 |  | 0.68 | 0.60 |  |  | 0.64 |
| 8. Could ask questions | 0.63 | 0.61 |  | 0.42 |  | 0.57 | 0.64 |  |  | 0.59 |
| 9. Checked understanding | 0.71 | 0.49 | 0.57 |  |  | 0.44 | 0.72 |  |  | 0.48 |
| 10. Birth position of choice | 0.53 | 0.72 | 0.44 |  |  | 0.67 | 0.48 |  |  | 0.77 |
| 11. Explain baby procedures | 0.63 | 0.60 | 0.30 | 0.41 |  | 0.58 | 0.70 |  |  | 0.51 |
| 12. Birth preferences respected | 0.52 | 0.73 | 0.36 |  |  | 0.72 | 0.54 |  |  | 0.71 |
| 13. Baby feeding choice respected | 0.58 | 0.66 |  | 0.32 |  | 0.66 | 0.56 |  |  | 0.69 |
| 14. Coercion | 0.13 | 0.98 |  |  | 0.47 | 0.78 | 0.10 |  |  | 0.99 |
| 15. Treated with respect | 0.76 | 0.42 |  | 0.63 |  | 0.38 |  | 0.75 |  | 0.44 |
| 16. Family respected | 0.79 | 0.38 | 0.70 |  |  | 0.33 |  | 0.80 |  | 0.36 |
| 17. Information confidential | 0.50 | 0.75 |  | 0.81 |  | 0.54 |  | 0.43 |  | 0.81 |
| 18. Privacy-covered | 0.46 | 0.79 |  | 0.41 |  | 0.71 |  | 0.44 |  | 0.80 |
| 19. Verbal abuse | 0.52 | 0.73 | 0.55 |  | 0.32 | 0.60 |  | 0.54 |  | 0.71 |
| 20. Physical abuse | 0.54 | 0.71 | 0.56 |  |  | 0.66 |  | 0.56 |  | 0.69 |
| 21. Discrimination | 0.74 | 0.45 | 0.74 |  |  | 0.37 |  | 0.81 |  | 0.34 |
| 22. Neglected | 0.61 | 0.63 | 0.59 |  | 0.35 | 0.49 |  | 0.62 |  | 0.61 |
| 23. Experience valued | 0.78 | 0.39 | 0.40 | 0.47 |  | 0.37 |  | 0.73 |  | 0.47 |
| 24. Customs respected | 0.60 | 0.64 | 0.71 |  |  | 0.53 |  | 0.63 |  | 0.60 |
| 25. Emotional well-being | 0.56 | 0.69 | 0.52 |  |  | 0.61 |  |  | 0.52 | 0.73 |
| 26. Pain management | 0.45 | 0.80 |  | 0.32 |  | 0.77 |  |  | 0.47 | 0.78 |
| 27. Took best care | 0.73 | 0.46 |  | 0.58 | 0.37 | 0.35 |  |  | 0.79 | 0.38 |
| 28. Trust | 0.73 | 0.46 |  | 0.75 |  | 0.33 |  |  | 0.78 | 0.40 |
| 29. Felt safe | 0.58 | 0.66 |  | 0.54 | 0.34 | 0.55 |  |  | 0.59 | 0.65 |
| 30. Companionship | 0.51 | 0.74 | 0.68 |  |  | 0.63 |  |  | 0.52 | 0.73 |
| 31. Timely response | 0.62 | 0.62 | 0.54 |  |  | 0.58 |  |  | 0.65 | 0.58 |
| 32. Believed about pain | 0.63 | 0.60 | 0.49 |  |  | 0.58 |  |  | 0.66 | 0.56 |
| 33. Support for baby feeding | 0.52 | 0.73 |  | 0.46 |  | 0.70 |  |  | 0.49 | 0.76 |
| 34. Comfortable birth environment | 0.75 | 0.43 | 0.72 |  |  | 0.38 |  |  | 0.73 | 0.47 |
| 35. Wait time | 0.50 | 0.75 | 0.35 |  |  | 0.73 |  |  | 0.50 | 0.75 |

Notes: F=Factor; CA=Communication & autonomy; DR=Dignity & respect; RS=Responsive & supportive

**Table 4:** Scale and sub-scale properties and distribution of scores (N=297)

|  | No. of items | Cronbach's alpha | Raw scores |  |  |  | Standardized score |  |  |  |
| --- | --- | --- | --- | --- | --- | --- | --- | --- | --- | --- |
|  |  |  | Mean | SD | Min | Max | Mean | SD | Min | Max |
| Full PCMC-US scale | 35 | 0.95 | 93.6 | 12.9 | 22.0 | 105.0 | 89.2 | 12.3 | 21.0 | 100.0 |
| Communication & autonomy | 14 | 0.87 | 37.1 | 5.2 | 11.0 | 42.0 | 88.4 | 12.3 | 26.2 | 100.0 |
| Dignity & respect | 10 | 0.87 | 27.7 | 3.6 | 7.0 | 30.0 | 92.4 | 12.1 | 23.3 | 100.0 |
| Responsive & supportive care | 11 | 0.87 | 28.8 | 4.8 | 4.0 | 33.0 | 87.2 | 14.6 | 12.1 | 100.0 |

To create summative scores for the full scale and sub-scales, the responses on the items for each are added. These scores are then standardized by dividing the mean score by the maximum possible scores ((e.g., 105 (35*3) for full scale, 42(14*3) for CA, 30(10*3) for DR, and 33(11*3) for RS)) and multiplying by 100. The standardized scores thus range from 0 to 100 for all the scores, where 0 is the worst PCMC and 100 is the best PCMC one can receive. The raw and standardized scores are shown in Table 4. The results show an average standardized PCMC score of 89. The highest sub-scale score is for Dignity and Respect (92) and the lowest score is for Communication and Autonomy (87).

The associations between the full PCMC-US scale and sub-scales and the related measures provided support for criterion validity. The regression of each of the sub-scales and the full scale on participants’ ratings of satisfaction with services, general quality ratings, and whether they would give birth in the same facility if they were to become pregnant again shows increasing PCMC is associated with higher odds of satisfaction and quality of care and intent to give birth in the same facility in the future (Table 5). The correlation coefficients between the PCMC-US scale and sub-scale scores and the MORi and MADM scale scores are all >0.5, with a correlation coefficient of 0.69 for CA and MADM and 0.63 for DR and MORi.

**Table 5:** Bivariate logistic and linear regression of scale scores on related measures to assess criterion validity (N=297)

|  | *Satisfied with care* |  |  | *Will birth in same place again* |  |  | *Rated quality of care as very good* |  |  | *MADM score* |  |  | *MORi score* |  |  |
| --- | --- | --- | --- | --- | --- | --- | --- | --- | --- | --- | --- | --- | --- | --- | --- |
| | *OR* | *95% CI* | | *OR* | *95% CI* | | *OR* | *95% CI* | | $\beta$ | *95% CI* | | $\beta$ | *95% CI* | |
| Full PCMC-US scale | 1.14*** | [1.09 | 1.18] | 1.09*** | [1.06 | 1.11] | 1.11*** | [1.07 | 1.14] | 0.35*** | [0.32 | 0.39] | 0.20*** | [0.17 | 0.22] |
| Communication & autonomy | 1.13*** | [1.09 | 1.17] | 1.08*** | [1.05 | 1.11] | 1.10*** | [1.07 | 1.13] | 0.34*** | [0.30 | 0.38] | 0.18*** | [0.15 | 0.21] |
| Dignity & respect | 1.10*** | [1.07 | 1.14] | 1.07*** | [1.04 | 1.09] | 1.10*** | [1.06 | 1.13] | 0.33*** | [0.28 | 0.37] | 0.20*** | [0.17 | 0.23] |
| Responsive & supportive care | 1.13*** | [1.09 | 1.17] | 1.08*** | [1.05 | 1.10] | 1.09*** | [1.06 | 1.12] | 0.28*** | [0.25 | 0.32] | 0.16*** | [0.13 | 0.18] |

Notes: \* p<0.05 \*\* p<0.01 \*\*\* p<0.001. Each row is a different bivariate model. MADM=Mothers Autonomy in Decision Making scale MORi= Mothers on Respect index. N=296 for MADM model;

## DISCUSSION

We used a rigorous, community-engaged approach to adapt the Person-Centered Maternity Care scale to reflect the experiences of people of color in the US. Through expert reviews and cognitive interviews, we developed 50-items that capture person-centered maternity care for women of color. We then used psychometric analysis and feedback from the CAB to reduce the items to a 35-item PCMC-US scale, which has three sub-scales measuring dignity and respect, communication and autonomy, and responsive and supportive care. The full scale and the sub-scales have good content, construct, and criterion validity with Cronbach’s alphas of 0.95 for the main scale and >0.8 for the sub-scales.

The final version of the PCMC-US scale ended up being similar to the original version (Afulani et al., 2017, 2018), with some subtle differences. Both versions have similar sub-scales, given the similar set of items and conceptual groupings using the WHO experience of care domains (Tunçalp et al., 2015). Although the wording of the questions resemble the original questions, the adaption process ensured that the exact wording of items in the new version are appropriate for the US context. The response options used in the original PCMC scale were maintained, as participants in the cognitive interviews found the frequency response format to be easy to respond to and did not think it should be changed. This response format leads to more variability in responses than binary yes/no responses. Acquiescence bias is also less of an issue than the agree/disagree format used in other scales (Holbrook, 2008).

The original PCMC scale has 30 items, compared to 35 items in the PCMC-US scale. This is due to several questions that were added during the adaptation process, leading to a set of 50 potential items, all of which were felt to be highly relevant by the cognitive interview participants and the CAB. The 35 items included in the PCMC-US scale are the result of our attempt to produce a parsimonious but comprehensive scale that incorporates feedback from the CAB and has high validity and reliability. Thus, although we excluded 15 items from the scale, these items could be included in surveys that are able to include a longer list of questions. In addition, we included the question on coercion despite its poor performance in factor analysis because of its importance to the experiences of women of color. The performance of this item should be reassessed in future studies.

The PCMC-US scale is different from other scales for measuring birthing people’s experience. First, to our knowledge it is the first available, validated person-centered care scale that centers the childbirth experiences of Black birthing people. Continuous engagement with members of the priority community ensured that the items in the scale capture their unique experiences (Altman et al., 2019, 2020; McLemore et al., 2018). The scale items were intentionally developed to include a mix of subjective questions to capture people’s subjective perceptions as well as more objective and actionable items that can inform quality improvement (Afulani, Aborigo, et al., 2019; Montagu et al., 2020). The PCMC-US scale is also among the few tools that measure experiences of person-centered care during childbirth in a comprehensive manner (Nilvér et al., 2017).

### Strengths and Limitations

A key strength of this study is the use of a community-based participatory approach embedded in standard instrument development methods. This helped ensure that the PCMC-US scale is relevant to people of color—particularly the Black community. Starting with a validated tool also provided a rigorous, theory-based foundation for the adaptation. A potential limitation is generalizability, given the relatively highly educated validation sample. This, however, is also a strength of the study, as it represents the views of a diverse group of Black people. Another limitation is the low representation of non-Black persons of color, which is likely because the survey was only administered in English. Plans for validation in a Spanish-speaking population are in place. The similarities between the original PCMC scale and the US version suggests that the scale may be applicable across different populations. Validations in new populations are however needed. Respondent burden due to the length of the scale may be a limitation. However, given the multidimensional nature of person-centered maternity care, the assessment of the relevance of the items included, and their psychometric performance, this is the most parsimonious version of the scale we are able to recommend at this time. The sub-scales can be used individually where necessary, but we recommend measuring all three domains to assess PCMC in a holistic manner: It takes about 10 minutes to answer all the questions. Shorter versions will be developed in future studies when data from more diverse samples are available, as was done for the low resource setting version (Afulani, Feeser, et al., 2019).

### Implications for Practice and/or Policy

Given the increasing documentation of the poor experience of people of color in health care settings, it is important that these experiences are documented in a systematic manner. In addition, there is the need to develop and evaluate interventions to improve the experiences of people of color in health care settings including during childbirth. These efforts require measures that center the experiences of the affected communities (K. Scott, 2019). The PCMC-US scale, together with ongoing efforts to measure obstetric racism (K. A. Scott & Davis, 2021), provide tools for these purposes.

## Conclusions

Using a community-engaged process, we adapted the PCMC scale that was initially developed in Kenya and India to make it relevant to the experiences of people of color in the US. The PCMC-US scale has high validity and reliability in a sample of predominantly black women. The scale will help drive as well as serve as an accountability tool in efforts to reduce disparities in pregnancy and birth outcomes.

## Data Availability

Data available from lead author on request

## Acknowledgements

We would like to thank all members of the California Preterm Birth Initiative Community Advisory Board, everyone who provided feedback at various phases of the scale development, and all the people who participated in the numerous stages of the project. We are also grateful to everyone who helped with recruitment and other members of our research team (Nadia Diamond-Smith and Nicholas Rubashkin at the University of California, San Francisco).

**Appendix A:**
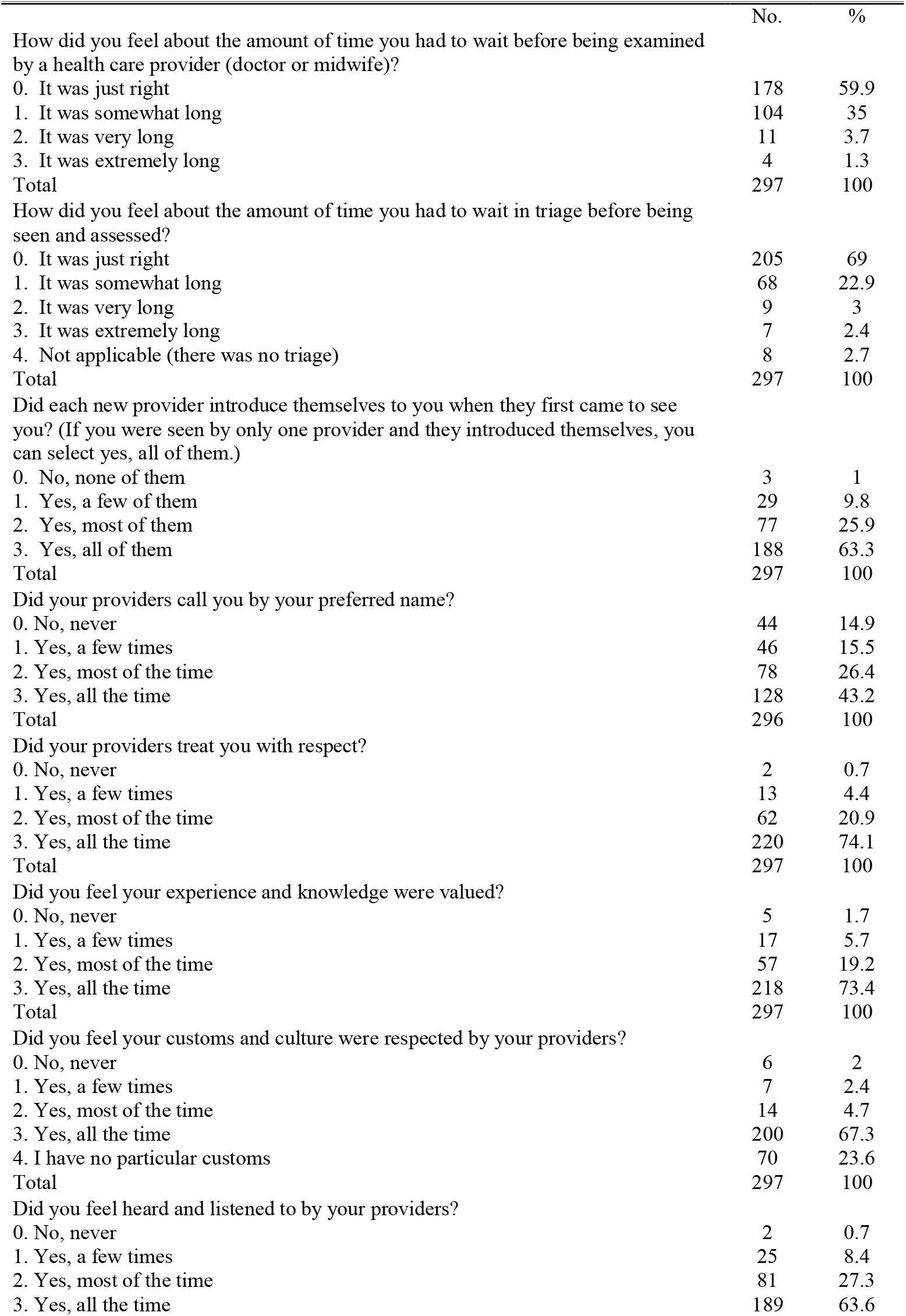
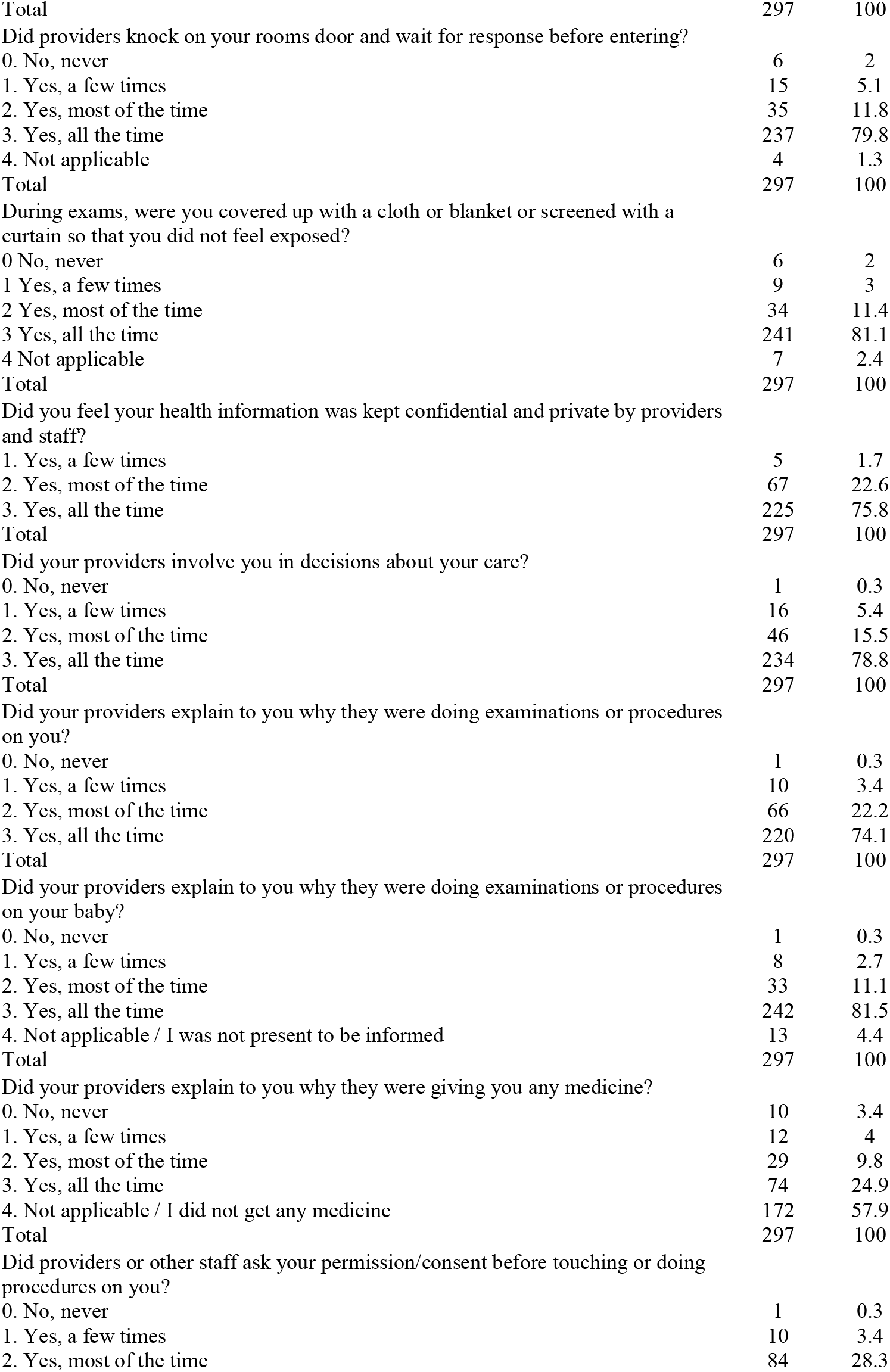
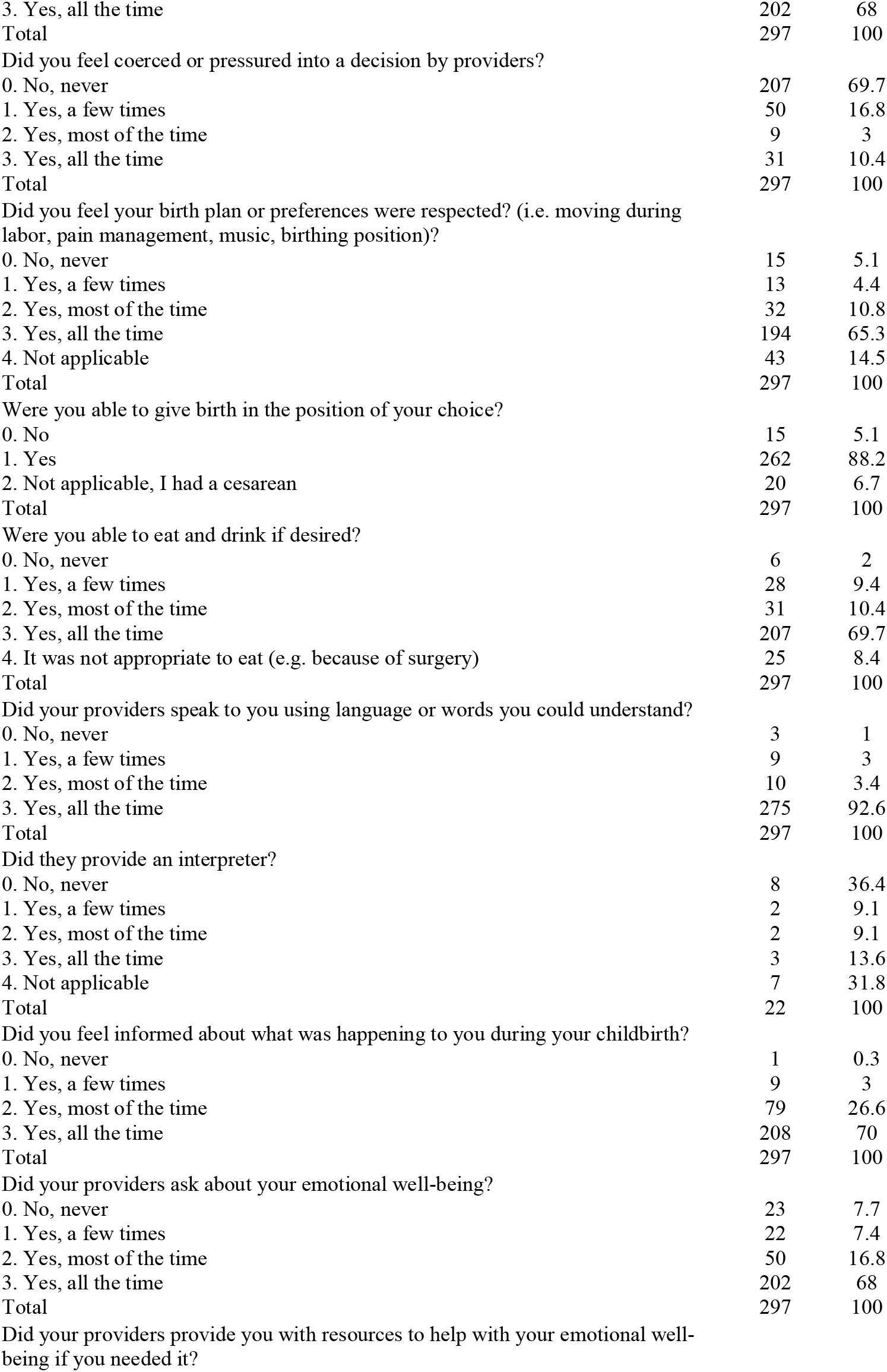
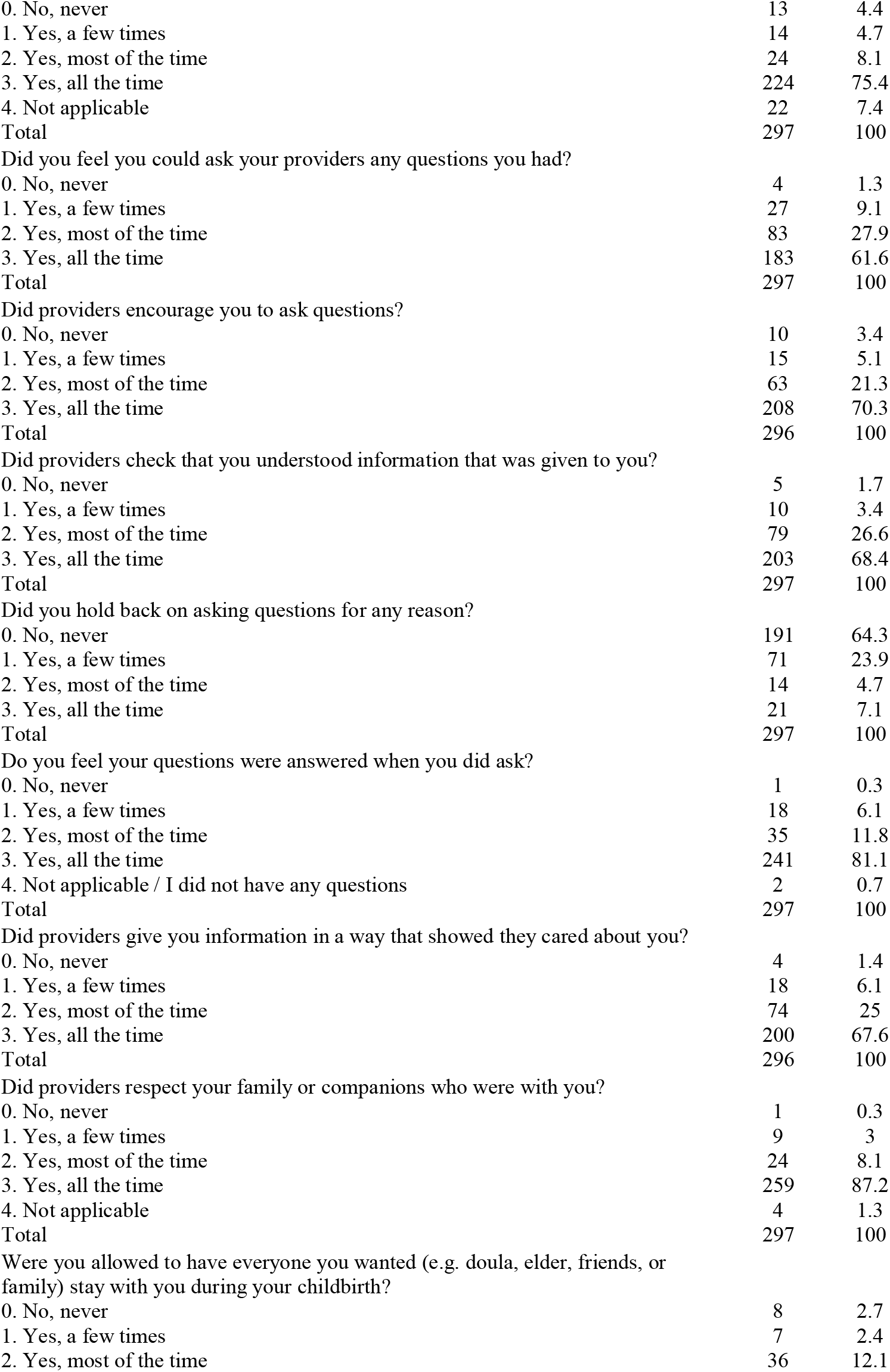
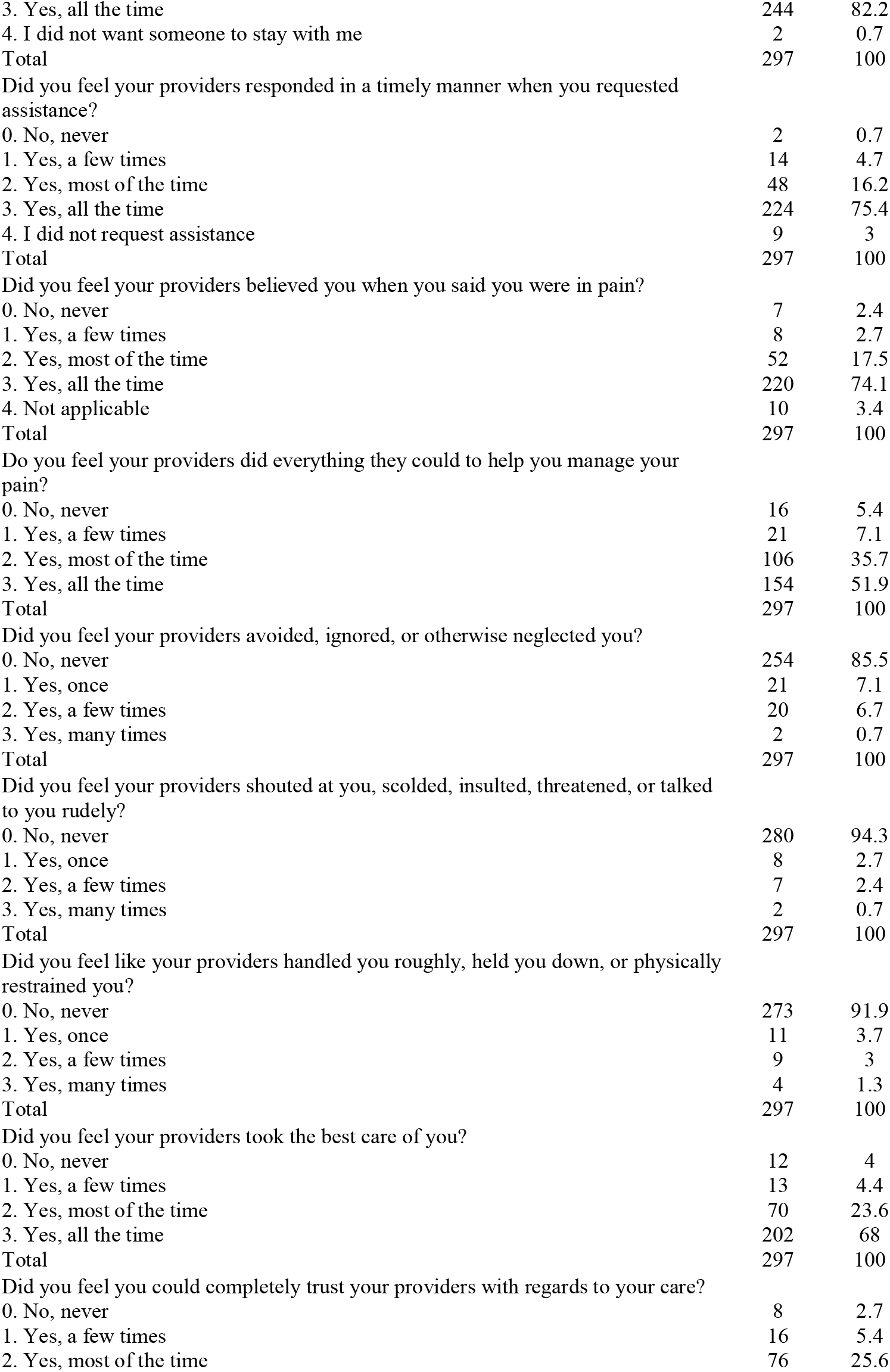
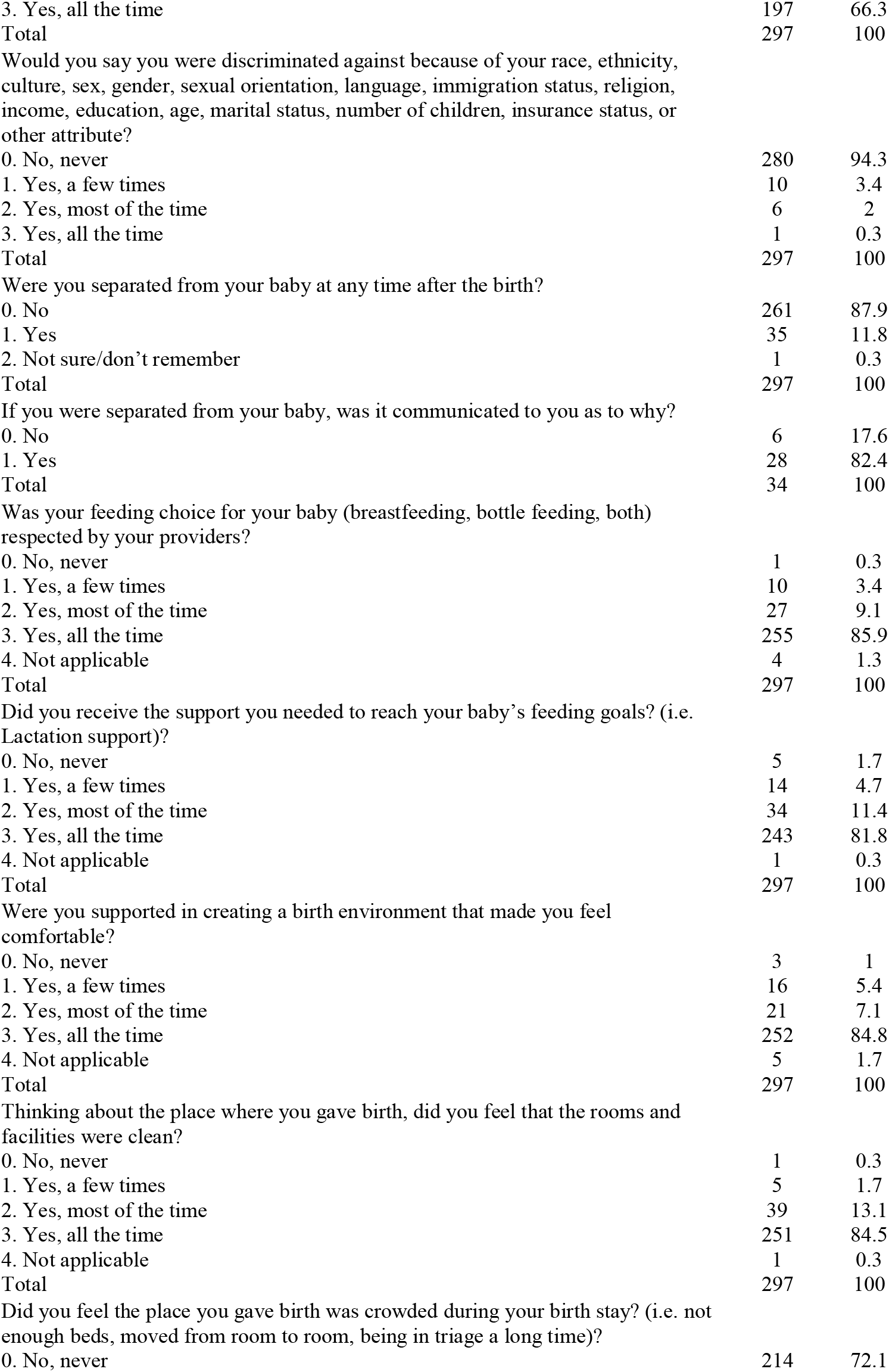
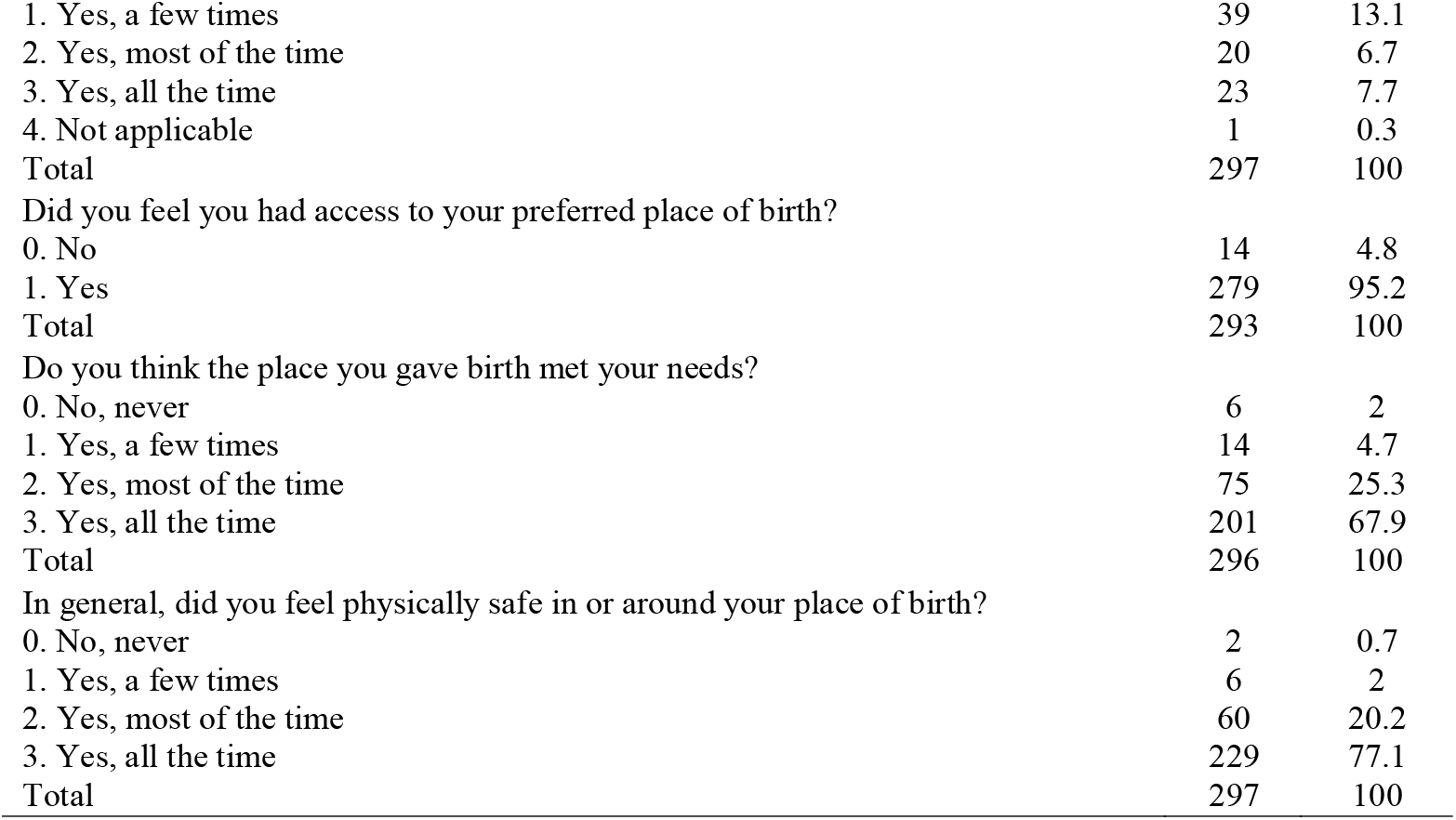
Distribution of Individual PCMC-US Items (N=297)

**Appendix B:** Results from Survey to Prioritize Items (N=14)

| No | Domain/Item | N | Notes |
| --- | --- | --- | --- |
| **Communication and Autonomy** |  |  |  |
| 1 | Did providers or other staff ask your permission/consent before touching or doing procedures or examinations on you? | 14 | High loading, High priority: retained |
| 2 | Did your providers explain to you why they were doing examinations or procedures on you? | 12 | High loading, High priority: retained |
| 3 | Did your providers explain to you why they were doing examinations or procedures on your baby | 10 | High loading, High priority: retained |
| 4 | Did your providers involve you in decisions about your care? | 10 | High loading, High priority: retained |
| 5 | Did you feel informed about what was happening to you during your childbirth? | 10 | High loading, High priority: retained |
| 6 | Did you feel coerced or pressured into a decision by providers? | 9 | High loading, High priority: retained |
| 7 | Did you feel you could ask your providers any questions you had? | 8 | High loading, High priority: retained |
| 8 | Did you feel heard and listened to by your providers? | 7 | High loading, High priority: retained |
| 9 | Did your providers speak to you using language or words you could understand? | 7 | High loading, High priority: retained |
| 10 | Did each new provider introduce themselves to you when they first came to see you? | 6 | High loading, Moderate priority: retained |
| 11 | Did providers check that you understood information that was given to you? | 6 | High loading, Moderate priority: retained |
| 12 | Was your feeding choice for your baby (breastfeeding, bottle) respected by providers? | 6 | High loading, Moderate priority: retained |
| 13 | Did your providers call you by your preferred name? | 5 | Low loading, Moderate priority: removed |
| 14 | Did you feel your birth plan or preferences were respected? (i.e. moving during labor, pain management, music, birthing position) | 5 | High loading, Moderate priority: retained |
| 15 | Were you able to give birth in the position of your choice? | 5 | High loading, Moderate priority: retained |
| 16 | Did your providers explain to you why they were giving medicine? | 4 | Low priority: removed |
| 17 | Did you hold back on asking questions for any reason? | 3 | Low priority: removed |
| 18 | Did providers encourage you to ask questions? | 3 | Low priority: removed |
| 19 | Do you feel your questions were answered when you did ask? | 2 | Low priority: removed |
| **Dignity and Respect** |  |  |  |
| 1 | Did your providers treat you with respect? | 14 | High loading, High priority: retained |
| 2 | Did providers respect your family or companions who were with you? | 14 | High loading, High priority: retained |
| 3 | Did you feel your providers shouted at you, scolded, insulted, threatened, or talked to you rudely? | 13 | High loading, High priority: retained |
| 4 | Would you say you were discriminated against because of your race, ethnicity, culture, sex, gender, sexual orientation, language, immigration status, religion, income, education, age, marital status, number of children, insurance status, or other attribute? | 13 | High loading, High priority: retained |
| 5 | Did you feel your providers avoided, ignored, or otherwise neglected you? | 12 | High loading, High priority: retained |
| 6 | Did you feel your customs and culture were respected by your providers? | 11 | High loading, High priority: retained |
| 7 | During exams, were you covered up with a cloth or blanket or screened with a curtain so that you did not feel exposed? | 11 | High loading, High priority: retained |
| 8 | Did you feel like your providers handled you roughly, held you down, or physically restrained you? | 8 | High loading, High priority: retained |
| 9 | Did you feel your experience and knowledge were valued? | 7 | High loading, High priority: retained |
| 10 | Did you feel your health information was kept confidential and private by providers? | 5 | High loading, moderate priority: retained |
| 11 | Did providers knock on your rooms door and wait for response before entering? | 3 | Low priority: removed |
| **Responsive and Supportive Care** |  |  |  |
| 1 | Did you feel you could completely trust your providers with regards to your care? | 12 | High loading, High priority: retained |
| 2 | Did you feel your providers responded in a timely manner when you requested assistance? | 11 | High loading, High priority: retained |
| 3 | Were you allowed to have everyone you wanted (e.g. doula, elder, friends, or family) stay with you during your childbirth? | 11 | High loading, High priority: retained |
| 4 | Did your providers ask about your emotional well-being? | 11 | High loading, High priority: retained |
| 5 | Do you feel your providers did everything they could to help you manage your pain? | 10 | High loading, High priority: retained |
| 6 | In general, did you feel physically safe in or around your place of birth? | 10 | High loading, High priority: retained |
| 7 | Did you feel your providers believed you when you said you were in pain? | 9 | High loading, High priority: retained |
| 8 | Did you receive the support you needed to reach feeding goals? (i.e. Lactation support) | 8 | High loading, High priority: retained |
| 9 | Were you supported in creating a birth environment that made you feel comfortable? | 8 | High loading, High priority: retained |
| 10 | How did you feel about the amount of time you had to wait before being examined by a health care provider (doctor or midwife)? | 7 | High loading, High priority: retained |
| 11 | Did you feel your providers took the best care of you? | 5 | High loading, Moderate priority: retained |
| 12 | Were you separated from your baby at any time after time after the birth? | 5 | Low loading, Moderate priority: removed |
| 13 | How did you feel about the time you waited in triage before being seen and assessed? | 4 | Low priority: removed |
| 14 | Did providers give you information in a way that showed they cared about you? | 4 | Low priority: removed |
| 15 | Thinking about the place where you gave birth, did you feel that the rooms and facilities were clean? | 4 | Low priority: removed |
| 16 | Did you feel the place you gave birth was crowded during your birth stay? (i.e., not enough beds, moved from room to room, being in triage a long time) | 4 | Low priority: removed |
| 17 | Did your providers provide you with resources to help with your emotional well-being if you needed it? | 3 | Low priority: removed |
| 18 | Do you think the place you gave birth met your needs? | 3 | Low priority: removed |
| 19 | Were you able to eat and drink if desired? | 2 | Low priority: removed |
| 20 | Did you feel you had access to your preferred place of birth? | 2 | Low priority: removed |

Notes: Considered high priority if selected by at least half of respondents (N>7) and low priority if selected by a third of respondents (<5).

